# Improving Frozen Kidney Biopsy Interpretation Using DUET-Generated Virtual PAS Stains

**DOI:** 10.1101/2025.11.11.25340018

**Authors:** Kuang-Yu Jen, Willy Ju, Srishti Seth, Ruby Mascareno, Ruben Carlos Renteria, Sarah Rezapourdamanab, Shalini Ramachandra, Jayla Hernandez, Sam Border, Ashna Aggarwal, Swikrity Upadhyay Baskota, Rajib Gupta, Alton B Farris, Fadi Salem, Priti Soin, Jarcy Zee, Pinaki Sarder, Farzad Fereidouni

## Abstract

Frozen section biopsies stained with Hematoxylin & Eosin (H&E) are standard for assessing donor kidneys but are often interpreted by general pathologists with limited renal expertise, and hindered by freezing artifacts and poor tissue morphology. There is currently no rapid, cost-effective, and easily deployable method for generating virtual special stains and quantifying clinically significant histopathological features on frozen section H&E slides.

This study evaluates the utility of DUET-generated virtual Periodic acid-Schiff (vPAS) stains produced from H&E-stained frozen kidney biopsies. DUET produces virtual PAS by combining pixel-registered brightfield and fluorescence images from the frozen section H&E-stained slide, overlaying extracted collagen masks to generate the virtual stain. Renal and general pathologists evaluated interstitial fibrosis/tubular atrophy (IF/TA), inflammation, and arteriosclerosis on 29 kidney biopsies, comparing their assessments using H&E images alone versus using both H&E and virtual PAS images.

Among general pathologists’ evaluations, ICC scores from the H&E to virtual PAS stain increased for IF/TA percentage, inflammation percentage, and arteriosclerosis number. For renal pathologists’ evaluations, ICC scores from the H&E to virtual PAS stain increased for the percentage of sclerotic glomeruli, IF/TA percentage, inflammation percentage, and number of arteriosclerotic lesions. No improvement in ICC score was observed for arteriolar hyalinosis in either pathologist’s group.

We have demonstrated that the use of frozen virtual PAS stains enables higher consistency among pathologist evaluations when compared to the use of frozen section H&E alone for various metrics used to assess donor kidney viability. This approach has the potential to reduce diagnostic variability, improve transplant decision-making, and optimize donor kidney utilization.

## Introduction

Deceased donor kidney biopsies are commonly performed as part of the overall assessment of organ quality for transplantation. Given that a rapid decision needs to be made as to whether the harvested kidney will be used for transplantation, frozen section analysis is standard of practice for evaluating these procurement biopsies in order to minimize cold ischemic time [1, 2].

However, frozen sections are plagued by numerous technical problems that render this modality highly inaccurate [3-5]. It is well known that frozen artifact greatly hinders the accurate interpretation of these biopsies [4]. Also, the frozen sections are stained with only hematoxylin and eosin (H&E). Special stains that greatly increase sensitivity and accuracy of diagnosis on renal biopsies that are routinely performed for formalin-fixed paraffin-embedded biopsies (gold standard) cannot be performed for frozen sections or be completed in the time frame that is needed by the transplant surgeons [6]. Special histochemical stains such as the periodic acid-Schiff (PAS) stain would significantly aid in assessing the degree of tubular atrophy, the presence of glomerular and vascular disease, and the percentage of glomerulosclerosis. However, its diagnostic utility is limited due to freezing artifacts and poor visualization of tissue morphology [7]. Although special stains can enhance interpretability, they are rarely used on frozen sections due to time-consuming protocols optimized for formalin-fixed paraffin-embedded (FFPE) tissue. To address these limitations, novel digital pathology approaches, including virtual staining, have emerged to extract clinically relevant features from frozen biopsies [8-12]. However, many of these methods rely solely on brightfield images or require advanced techniques such as confocal or stimulated Raman scattering (SRS) microscopy, making them expensive and impractical for routine clinical use [13-16].

To overcome these challenges, our lab developed DUal-mode Emission and Transmission (DUET) microscopy, a novel technique that enables collagen extraction directly from H&E-stained frozen sections. DUET allows generation of virtual PAS images without the need for additional tissue sections or specialized imaging platforms [17].

In this study, we evaluate the effectiveness of DUET-generated virtual PAS stains in enhancing pathologists’ assessment of donor kidney viability. We hypothesize that these virtual stains will help both general and renal pathologists make more accurate and consistent evaluations under time constraints, addressing the variability and limitations of current frozen section workflows. We demonstrate that incorporating virtual PAS information improves inter-pathologist agreement, scoring consistency, and diagnostic reliability compared to using H&E alone. With further validation and clinical translation, this approach has the potential to reduce organ discard rates and ease the diagnostic burden on on-call pathologists during transplantation procedures.

## Materials & Methods

### Image Acquisition using DUET

DUET images are generated by scanning pixel-registered brightfield and fluorescence images from frozen section hematoxylin and eosin (H&E) stained kidney biopsy slides. For fluorescence images, we employ epifluorescent excitation of H&E-stained tissue using a 405nm LED (LZ1-00UB00-LED Engin) directed towards the sample using a broadband dichroic (Di03-R405-t1-25 x 36, Semrock) and focused using a 10X objective (Nikon, Plan Apo, 0.45 NA). A 9-megapixel scientific-grade CCD color camera (Ximea, MD091CU-SY) connected via a 200mm tube lens (Thorlabs ILT 200) was used to capture images [17].

Slides were mounted on a custom-designed holder, and three Zaber motorized stages were used to perform XY(X-ASR050B050B) scanning and Z-axis (X-DMQ12L) focus adjustment. Image acquisition and processing were controlled through custom software developed in a .NET environment.

### Image Processing – Virtual Stain Generation

Fluorescence images were normalized to the total intensity at the pixel level, and a few regions were annotated for collagen and cytoplasm on each slide. An average of 20 points was used to unmix the collagen signal using linear discriminant analysis [18]. The virtual PAS images were created by using the matrix illustrated in Figure 1. The virtual stain color profiles were adjusted to match standard PAS stain colors, typically shades of magenta and purple (Figure 1).

**Figure 1.**
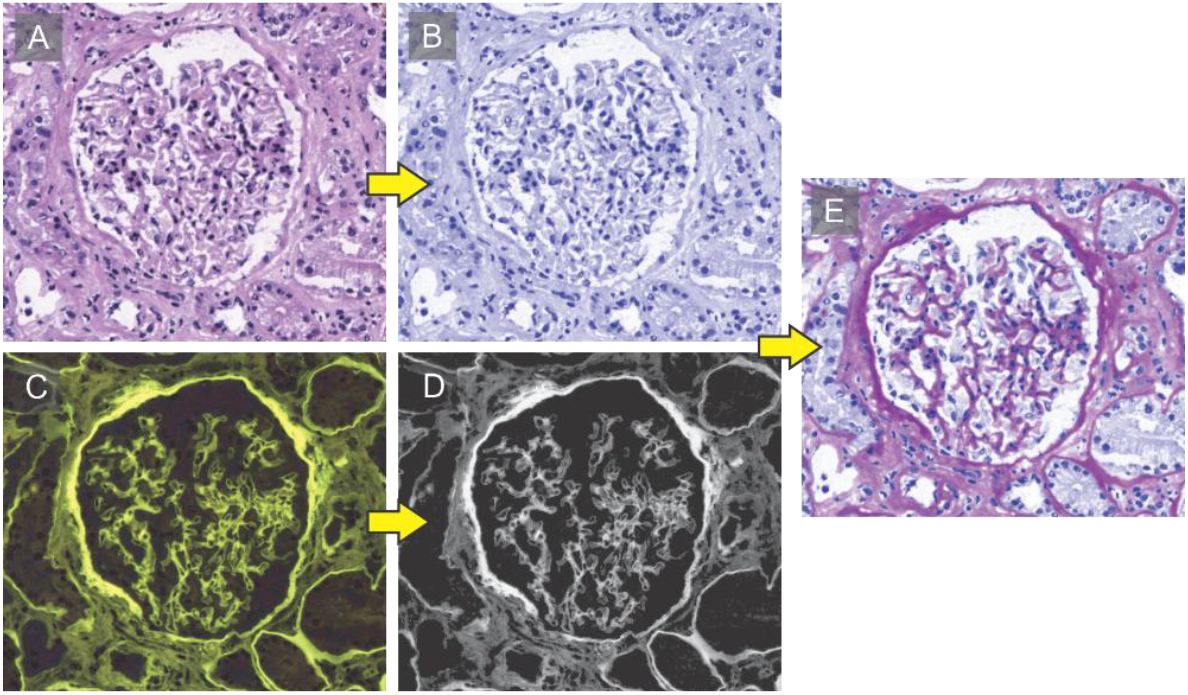
Panel showing visual workflow of methods used in DUET scanning of brightfield, fluorescence, extraction, and virtually generated PAS from the same frozen section kidney H&E slide. Pixel-matched brightfield and fluorescence images were acquired using a 10X objective. Collagen is extracted using a semi-automated method and overlaid on the augmented brightfield image to generate a virtual PAS stain.

### Pathologist Evaluation

Twenty-nine frozen kidney sections were scanned using DUET to generate Brightfield H&E and corresponding virtual PAS whole slide images (WSIs). Four renal pathologists and three general surgical pathologists scored these images using the following criteria:

The evaluation includes counting all glomeruli, including globally sclerotic ones, with segmental sclerosis noted separately. Interstitial fibrosis and tubular atrophy (IF/TA) are quantified to the nearest 5%, with <5% indicating minimal or absent findings. Inflammation is similarly estimated to the nearest 5%; use <5% for minimal or absent; typically within areas of IF/TA. Arteriosclerosis is assessed based on the degree of intimal thickening in arteries, rated as absent, mild (intimal thickening less than the thickness of media), moderate (intimal thickening equal to or up to twice the thickness of media), or severe (intimal thickening greater than twice the thickness of media). Hyaline arteriolosclerosis is scored based on arterioles only, from absent, mild (rate arterioles with a small amount of hyalin), moderate (scattered arterioles with moderate amounts of hyalin), and severe (frequent arterioles with prominent amounts of hyalin; often with luminal narrowing) [19]. The pathologists first assessed only the H&E WSIs and scored the features based on the criteria specified above. Then, following a two-week washout period, the pathologists reviewed the cases with both H&E and virtual PAS WSIs.

### Statistical Analysis

Since each sample was rated twice per rater for a total of 14 observations per sample, the intraclass correlation coefficient (ICC), which measures variability within samples for repeated observations, was calculated overall and within subgroups. Inter-rater ICCs were calculated to estimate agreement across pathologists for each stain, and intra-rater ICCs were calculated to estimate agreement across stains for each pathologist. The ICC was calculated as a ratio of between-sample variability and the sum of between- and within-sample variability based on a random effects model, so it is bounded by zero. A two-way random effects model was used for overall intra-rater ICCs, i.e., ICC(2,1), and a one-way random effects model was used for all other ICCs, i.e., ICC(1,1). 95% parametrically bootstrapped confidence intervals were calculated and reported for each ICC using the bootMer function in the lme4 R package with 1000 iterations.

To test if the differences between inter-rater ICCs by stain type within each pathologist category (overall, general, and renal) were statistically significant, permutation tests were performed. Stain type labels for each case were randomly permuted over 10,000 repetitions and a two-sided p-value was computed by comparing the observed difference in ICCs to the permutation-based distribution. We applied a Benjamini-Hochberg correction for multiple comparisons, a common method to control false discovery rate (FDR), and the FDR-adjusted p-values (“q-values”) were computed.

## Results

Figure 2 demonstrates a visual comparison between a standard frozen H&E slide and a DUET-generated virtual PAS image from the same tissue section. In the H&E images, collagen within the glomeruli and basement membranes shows minimal contrast, making the fibrotic regions less visually distinct from healthy areas in the tissue. However, these features are readily noticeable in the virtual PAS stain and are more visually contrasting compared to the surrounding tubules. This enhanced detail allows for more straightforward identification and interpretability of these structures.

**Figure 2.**
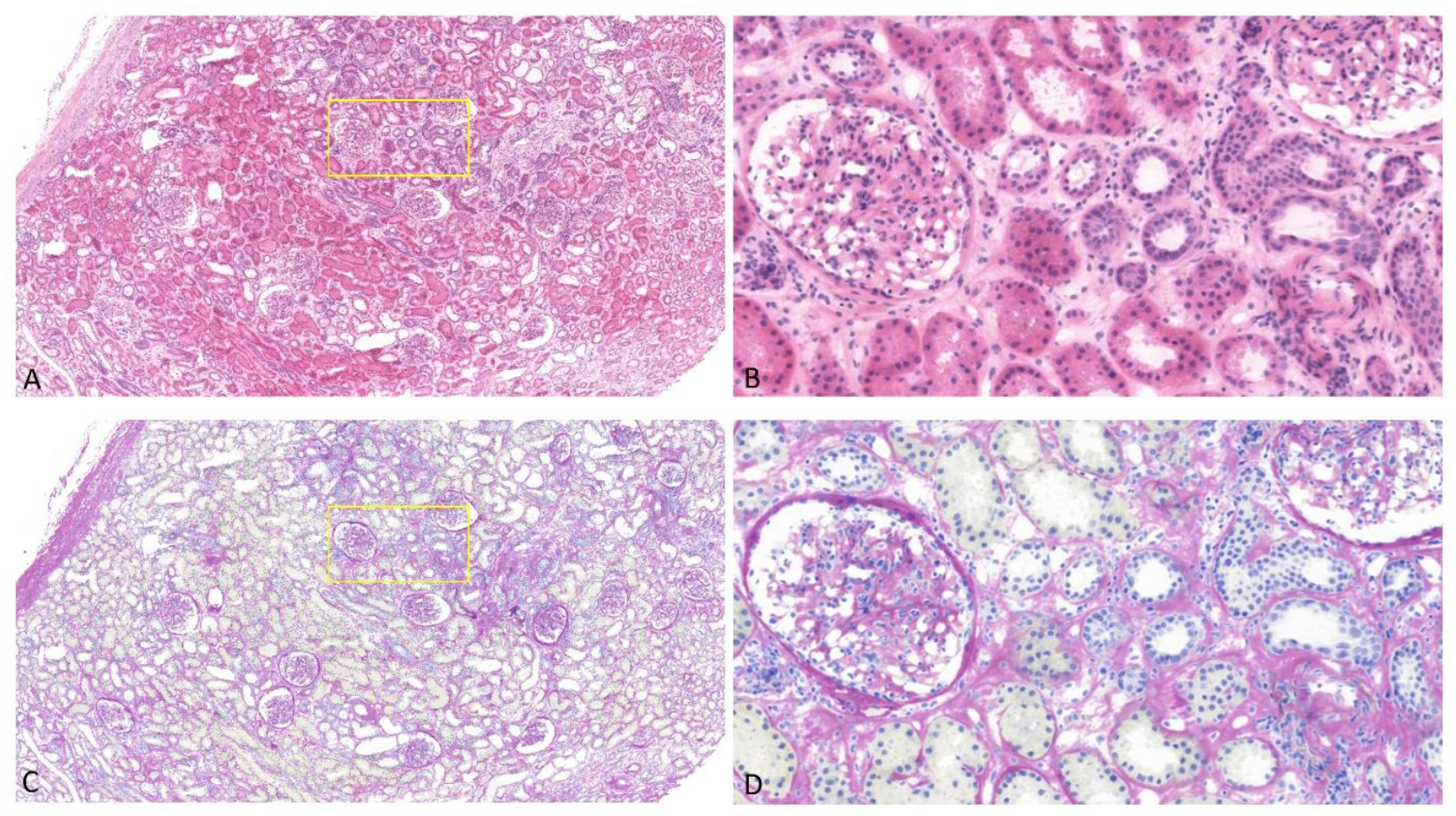
Visual comparison of frozen section H&E and virtual PAS scan of frozen kidney biopsy. Yellow box highlights zoomed in regions of interest A) H&E scan, B) Zoomed in area of H&E highlighting glomeruli and tubules, C) Pixel-matched DUET-generated Virtual PAS, D) Zoomed in area of Virtual PAS highlighting glomeruli and tubules.

We observed an improvement in inter-pathologist reliability among both general and renal pathologists across several key histological features. For general pathologists’ evaluations, ICC scores from the H&E to virtual PAS stain increased for IF/TA percentage (0.103 to 0.722, q-value = 0.0045), inflammation percentage (0.208 to 0.310, q-value = 0.55), and arteriosclerosis number (0.033 to 0.428, q-value = 0.054). For renal pathologists’ evaluations, ICC scores from the H&E to virtual PAS stain increased for percent sclerotic glomeruli (0.794 to 0.877, q-value = 0.038), IF/TA percentage (0.201 to 0.435, q-value = 0.13), inflammation percentage (0.031 to 0.343, q-value = 0.13), and arteriosclerosis number (0.581 to 0.615 q-value = 0.91). No improvement in ICC score was observed for arteriolar hyalinosis in either pathologists’ group, signifying consistent difficulty in evaluating this tissue feature across both stains (Table 1, Figure 3).

**Table 1.** Inter-rater (across all pathologists, across general pathologists, and across renal pathologists) intraclass correlation coefficient (ICC) for scores by stain, with 95% confidence intervals.

| Stain Type | ICC (95% CI) |  |  |  |  |
| --- | --- | --- | --- | --- | --- |
|  | Sclerotic Glom % | IF / TA % | Inflammation % | Arteriosclerosis # | Arteriolar Hyalinosis # |
| H&E |  |  |  |  |  |
| Overall | 0.726 (0.586, 0.821) | 0.177 (0.044, 0.302) | 0.107 (0, 0.228) | 0.358 (0.196, 0.505) | 0.203 (0.061, 0.344) |
| General | 0.668 (0.451, 0.794) | 0.103 (0, 0.324) | 0.208 (0, 0.433) | 0.033 (0, 0.269) | 0.128 (0, 0.364) |
| Renal | 0.784 (0.639, 0.869) | 0.201 (0.008, 0.378) | 0.031 (0, 0.195) | 0.581 (0.375, 0.720) | 0.358 (0.139, 0.520) |
| PAS |  |  |  |  |  |
| Overall | 0.784 (0.658, 0.862) | 0.515 (0.341, 0.644) | 0.337 (0.166, 0.485) | 0.499 (0.317, 0.639) | 0.137 (0.019, 0.272) |
| General | 0.635 (0.415, 0.777) | 0.722 (0.540, 0.828) | 0.310 (0.032, 0.526) | 0.428 (0.178, 0.610) | 0 (0, 0.227) |
| Renal | 0.877 (0.788, 0.927) | 0.435 (0.230, 0.612) | 0.343 (0.136, 0.524) | 0.615 (0.438, 0.755) | 0.339 (0.122, 0.513) |

**Figure 3.**
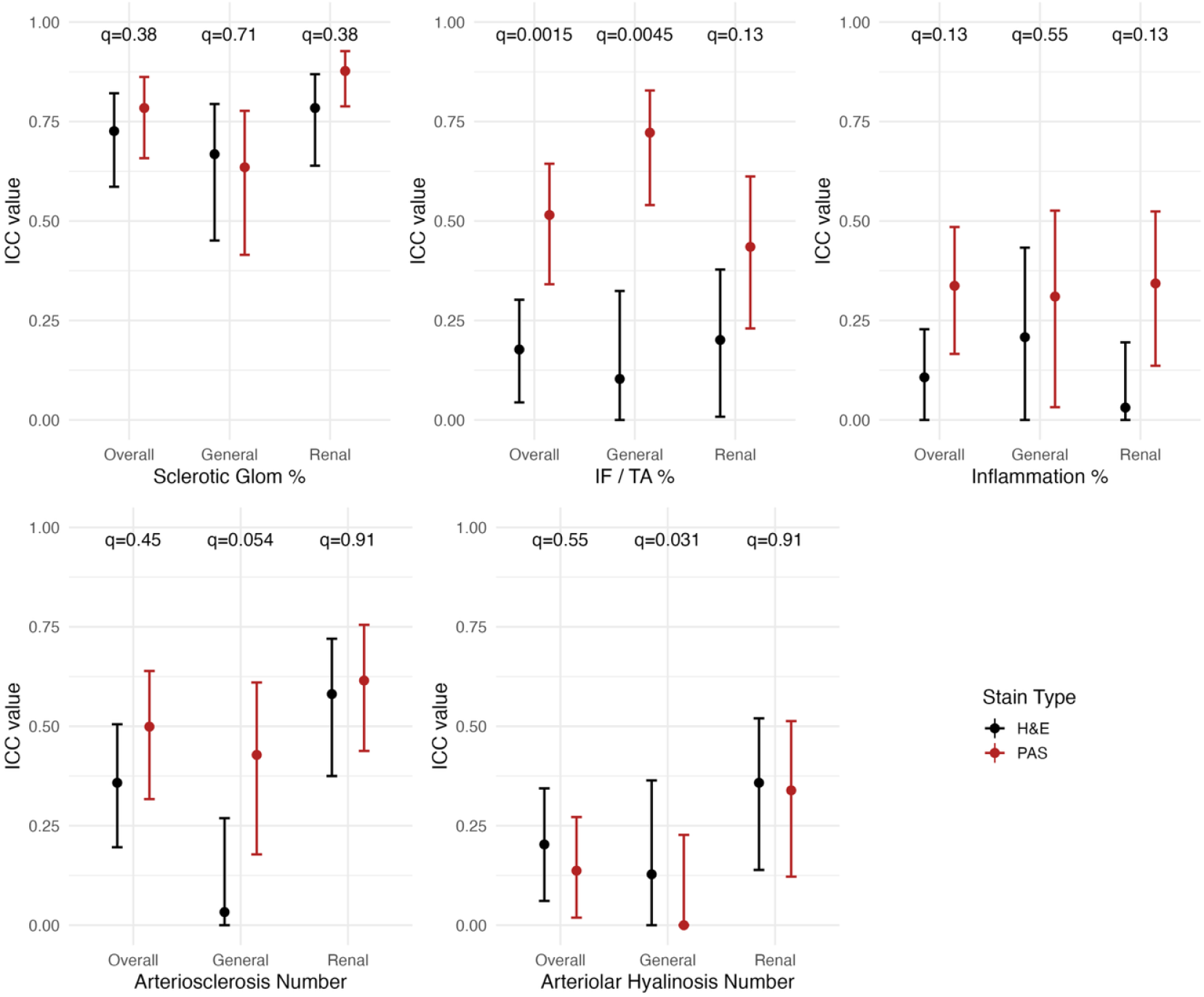
Inter-rater intraclass correlation coefficient (ICC) by pathologist type and overall, for each stain, with 95% confidence interval bars and false discovery rate adjusted p-values (“q-values”).

DUET showcases various strengths in identifying specific histologic details such as interstitial fibrosis, tubular atrophy (IF/TA), glomerulosclerosis, and nodules through the virtual PAS stain. While these features are present on the standard frozen H&E stain, it is difficult to clearly differentiate the fibrous formation between the tubules from the inside of the tubules (Figure 4A). The shriveled shape of the tubules from the frozen H&E makes visualizing IF/TA challenging, as the frozen artifacts make the details more blurred. Moreover, the severity of IF/TA is much more subtle in the H&E stain compared to the virtual PAS stain, where the stark purple contrast between the tubules and the thickening of the edges of the tubules is much more apparent (Figure 4B). These details are more similar to the IF/TA visualization from a real PAS stain on FFPE kidney tissue (Figure 4C).

**Figure 4.**
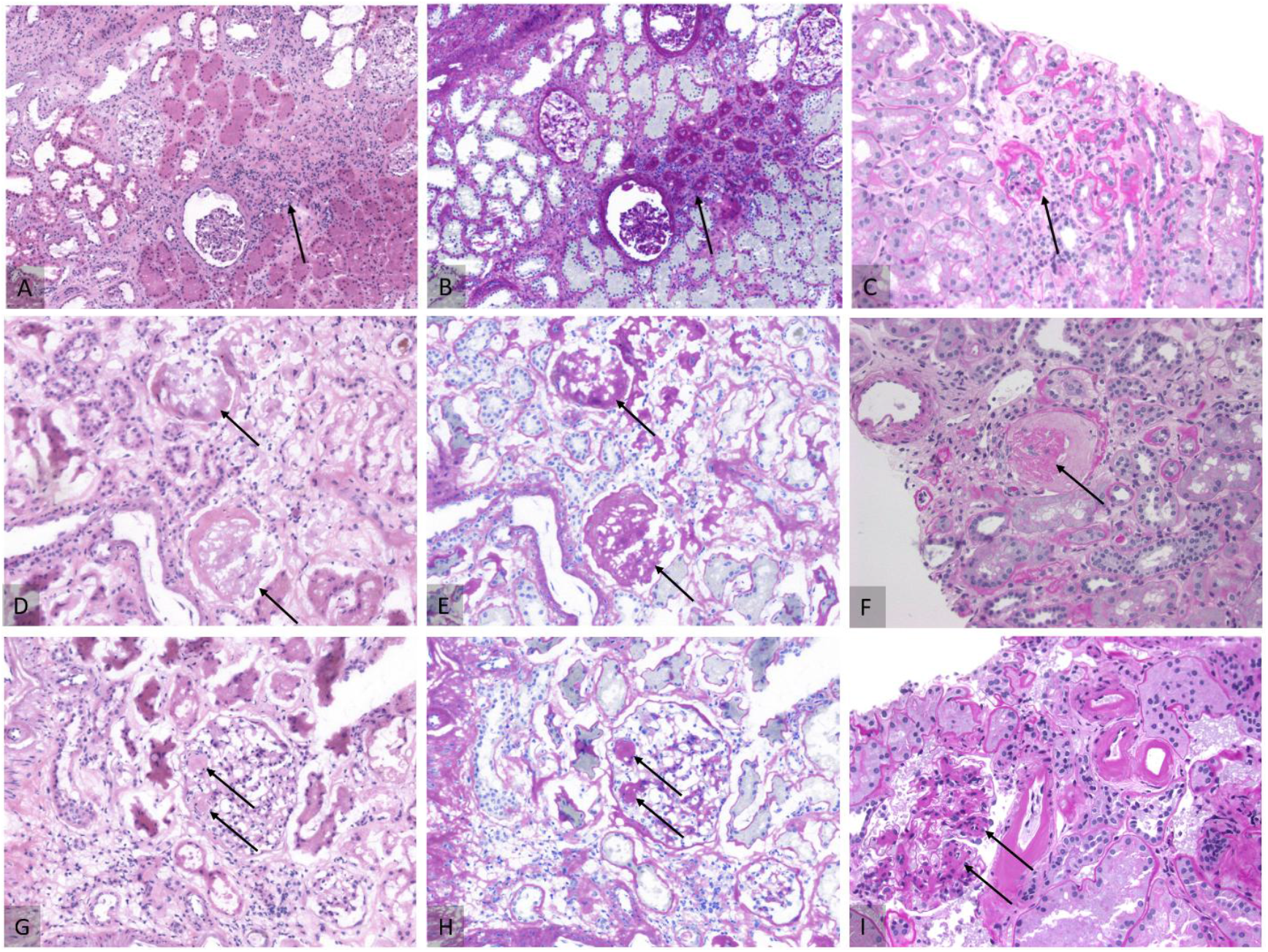
Strengths in DUET virtual PAS in interstitial fibrosis/tubular atrophy (IF/TA), glomerulosclerosis, nodules detection (marked by arrow). A) H&E scan of IFTA B) DUET-generated virtual PAS scan comparison of IFTA C) Real PAS scan of IFTA D) H&E scan of glomerulosclerosis E) DUET-generated virtual PAS scan comparison of glomerulosclerosis F) Real PAS scan of glomerulosclerosis G) H&E scan of nodules H) DUET-generated virtual PAS scan comparison of nodules I) Real PAS Scan of nodules.

Similarly, the sclerotic glomeruli and nodules are close in color to the nearby tubules on the H&E stain, thus making it arduous to distinguish these features within a short period of time (Figure 4D, G). The virtual PAS stain, however, can clearly illustrate the difference between a sclerotic and healthy glomerulus without studying the biopsy too closely. The sclerotic glomerulus is stained completely purple, whereas the healthy glomerulus still retains its nuclear and capillary detail. The virtual PAS stain also shows the nodules as distinct circular, purple structures that contrast with the rest of the glomerulus and nearby gray colored tubules (Figure 4E, H). Images of a sclerotic glomerulus and mesangial nodule in FFPE kidney tissue highlight the similarities between the virtual and real PAS stains (Figure 4F, I).

Certain tissue features, such as fibrin and arteriolar hyalinosis, were not clearly delineated in the virtual PAS images generated by DUET microscopy. In the H&E image, fibrin appears as saturated red deposits within the glomerulus (Figure 5A). However, in the corresponding virtual PAS image, this signal is absent, and the fibrin blends in with the gray counterstain, resembling the luminal content of tubules (Figure 5B). In contrast, the real PAS stain also does not highlight fibrin (Figure 5C). A similar finding was observed for arteriolar hyalinosis. In the H&E image, hyalinosis appears as a small, eosinophilic, circular ring adjacent to the glomerulus (Figure 5D). The virtual PAS image does not distinguish this structure, and it blends into the gray background (Figure 5E). The real PAS stain of FFPE kidney tissue, however, highlights this feature with a magenta signal (Figure 5F).

## Discussion

In this work, we demonstrated that a frozen section virtual PAS stain shows higher consistency (as measured by two-way ICC) among pathologist evaluations when compared to a frozen section H&E stain for various metrics used to assess donor kidney viability. Frozen biopsies inherently result in lower H&E quality due to frozen artifacts and tissue degradation [20]. Despite this, the virtual PAS stain still shows the color contrast between fibrous areas and cytoplasm. This virtual stain allows for easier visualization of IF/TA, glomerulosclerosis, and nodules, as ICC scores for general and renal pathologists increased when comparing the virtual stain to a standard H&E stain for frozen wedge kidney biopsies. The virtual stains could not delineate fibrin from the fluorescence of the frozen H&E stain. However, this is consistent with a real PAS stain in this regard, as PAS cannot detect fibrin either [21]. Arteriolar hyalinosis is another tissue feature that could not be detected from the virtual PAS stain. This would be magenta-colored in a real PAS stain [21], distinguishing its signal from the counterstain. Because hyalinosis is not made of collagen [22], it does not fluoresce under DUET microscopy’s fluorescent mode, and would not be turned into a positive signal, thus resulting in the gray color of the counter stain.

The advantages of the virtual PAS stain become particularly significant when considered in the context of the current challenges associated with a frozen section setting. The standard workflow for creating frozen biopsies from donor organs results in poor-quality biopsies, making it exceedingly difficult for on-call pathologists to interpret tissue features. There is also a large variety in the composition and protocol of H&E reagents used for staining, resulting in biopsies that look visually different from one another [23]. Despite being from the same institution, samples in this study were either under-stained or over-stained, resulting in different color profiles of the H&E stain. Additionally, there is subjective interpretability among different pathologists [24] and across different medical centers [25], proving the need for a more standardized evaluation of biopsies. The virtual PAS stain used in this study, by comparison, shows a more similar color profile across samples that otherwise have drastically different H&E-stained results.

This particular stain can also increase accessibility to information that is typically not possible to replicate in standard clinical workflow. Because the virtual PAS stain is created after the frozen biopsy has been collected and stained, this method does not disrupt the routine staining protocol, but rather supplements the existing H&E slide for the reviewing pathologist to make an informed decision.

Previous literature has also cited benefits for virtual stains to reduce labor costs and increase efficiency of the existing histology workflow [26]. Though this virtual PAS stain could theoretically be applied to FFPE biopsies that require special stains for further visualization, the primary goal of this project is to introduce a novel, non-disruptive method for evaluating frozen biopsies, particularly in the context of donor kidney assessment, where pathologists often face time pressure and poor tissue quality. Future studies will explore the automation of the collagen training to create masks for a more standardized methodology. We hope to create a more adaptive workflow that can account for the subjective nature of frozen H&E quality and the color profile of stains. The workflow proposed here to create the virtual PAS stain is intended to supplement, rather than replace, the frozen H&E stain. By evaluating both the frozen H&E and virtual PAS stains together, we hope that on-call pathologists will be able to use the strengths of each stain to gather more information about viability within the limited time available for transplant decision-making.

To date, there has been no method proposed to generate special stains directly from frozen section biopsies. In this paper, we have outlined a novel, feasible method to leverage the existing fluorescence from eosin to create a virtual PAS stain from existing frozen H&E biopsies. Virtual stains are especially important for accurate analysis of biopsies, potentially reducing the discard rate of otherwise viable organs. We hope that a virtual PAS stain will alleviate the burden on pathologists who may not have extensive renal pathology experience and are expected to decide whether or not to approve transplantation of a kidney.

## Data Availability

All data produced in the present study are available upon reasonable request to the authors

## Data availability

Data underlying the results presented in this paper are not publicly available at this time but may be obtained from the authors upon reasonable request.

## References

1. Randhawa, P., Role of donor kidney biopsies in renal transplantation. Transplantation, 2001. 71(10): p. 1361–1365.

2. Novis, D.A. and R.J. Zarbo, Interinstitutional comparison of frozen section turnaround time. Archives of pathology & laboratory medicine, 1997. 121(6): p. 559.

3. Azancot, M.A., et al., The reproducibility and predictive value on outcome of renal biopsies from expanded criteria donors. Kidney Int, 2014. 85(5): p. 1161–8.

4. Liapis, H., et al., Banff histopathological consensus criteria for preimplantation kidney biopsies. American Journal of Transplantation, 2017. 17(1): p. 140–150.

5. Wang, C., et al., The donor kidney biopsy and its implications in predicting graft outcomes: a systematic review. American Journal of Transplantation, 2015. 15(7): p. 1903–1914.

6. Walker, P.D., T. Cavallo, and S.M. Bonsib, Practice guidelines for the renal biopsy. Mod Pathol, 2004. 17(12): p. 1555–63.

7. Goumenos, D.S., et al., The prognostic value of frozen section preimplantation graft biopsy in the outcome of renal transplantation. Renal failure, 2010. 32(4): p. 434–439.

8. Marsh, J.N., et al., Deep learning global glomerulosclerosis in transplant kidney frozen sections. IEEE transactions on medical imaging, 2018. 37(12): p. 2718–2728.

9. Bevilacqua, V., et al., An innovative neural network framework to classify blood vessels and tubules based on Haralick features evaluated in histological images of kidney biopsy. Neurocomputing, 2017. 228: p. 143–153.

10. Salvi, M., et al., Automated assessment of glomerulosclerosis and tubular atrophy using deep learning. Computerized Medical Imaging and Graphics, 2021. 90: p. 101930.

11. Gorman, B.G., M.A. Lifson, and N.Y. Vidal, Artificial intelligence and frozen section histopathology: a systematic review. Journal of Cutaneous Pathology, 2023. 50(9): p. 852–859.

12. Ozyoruk, K.B., et al., Deep learning-based frozen section to FFPE translation. arXiv preprint arXiv:2107.11786, 2021.

13. Pillar, N., et al., Virtual staining of non-fixed tissue histology. Modern Pathology, 2024: p. 100444.

14. Falahkheirkhah, K., et al., Accelerating cancer histopathology workflows with chemical imaging and machine learning. Cancer Research Communications, 2023. 3(9): p. 1875–1887.

15. Krishnamurthy, S., et al., Confocal fluorescence microscopy platform suitable for rapid evaluation of small fragments of tissue in surgical pathology practice. Archives of Pathology & Laboratory Medicine, 2019. 143(3): p. 305–313.

16. Liu, J., et al., Nondestructive diagnosis of kidney cancer on 18-gauge core needle renal biopsy using dual-color fluorescence structured illumination microscopy. Urology, 2016. 98: p. 195–199.

17. Fereidouni, F., et al., Dual-mode emission and transmission microscopy for virtual histochemistry using hematoxylin-and eosin-stained tissue sections. Biomedical optics express, 2019. 10(12): p. 6516–6530.

18. Fereidouni, F., A.N. Bader, and H.C. Gerritsen, Spectral phasor analysis allows rapid and reliable unmixing of fluorescence microscopy spectral images. Optics express, 2012. 20(12): p. 12729–12741.

19. Roufosse, C., et al., A 2018 Reference Guide to the Banff Classification of Renal Allograft Pathology. Transplantation, 2018.

20. Kumar, K., D.C. Shetty, and M. Dua, Biopsy and tissue processing artifacts in oral mucosal tissues. Int J Head Neck Surg, 2012. 3(2): p. 92–8.

21. Agarwal, S., S. Sethi, and A. Dinda, Basics of kidney biopsy: A nephrologist’s perspective. Indian journal of nephrology, 2013. 23(4): p. 243–252.

22. Barbosa, G.S.B., et al., Vascular injury in glomerulopathies: the role of the endothelium. Frontiers in Nephrology, 2024. 4: p. 1396588.

23. Tosta, T.A.A., et al., Computational normalization of H&E-stained histological images: Progress, challenges and future potential. Artificial intelligence in medicine, 2019. 95: p. 118–132.

24. Haas, M., Donor kidney biopsies: pathology matters, and so does the pathologist. Kidney international, 2014. 85(5): p. 1016–1019.

25. Reeve, J., et al., Diagnosing rejection in renal transplants: a comparison of molecular-and histopathology-based approaches. American journal of transplantation, 2009. 9(8): p. 1802–1810.

26. Li, Y., et al., Virtual histological staining of unlabeled autopsy tissue. Nature Communications, 2024. 15(1): p. 1684.

